# Antimicrobial resistance patterns of *Escherichia coli* isolated from raw cow milk and, from clinical specimens in a tertiary institution, Uganda -A cross sectional study

**DOI:** 10.1101/2025.03.06.25323542

**Authors:** Jacob Michael Othieno, Anastacia Sebbowa N, Abel Wembabazi, Brain Martin Odhiambo, Winnie Nalwanga, Amusa Wamawobe, Sabrina Bakeera-Kitaka, Beatrice Achan

## Abstract

**Background:** The drastic rise of antimicrobial resistance to alarming states is one of the major concerns of global health today. The overuse of antibiotics in animal husbandry has tremendously contributed to increased emergence of antimicrobial resistant bacteria. These resistant strains from animals can be transferred to humans through the food chain like raw milk which is a potential source of food-borne bacterial pathogens including *E. coli*.

**Objective:** The objective of this study was to compare the patterns of antibacterial resistance of the *E. coli* isolates of animal origin from raw milk samples and *E. coli* isolates obtained from clinical specimens in the Clinical Microbiology Laboratory of Makerere University.

**Methodology:** A cross-sectional study with purposive sampling was conducted between January and March 2024, on N= 124 raw cow milk samples obtained from diary shops in Kawempe division and 13 clinical samples obtained from the Clinical Microbiology Laboratory of Makerere University. Samples were initially enriched in nutrient broth, sub-cultured MacConkey agar and biochemical tests were used to identify samples with *E. coli* from which prevalencewas determined. The antibacterial resistance pattern of *E. coli* against the selected antibiotics was determined using the disk diffusion susceptibility method.

**Results:** Out of the 124 milk samples (77 from cans and 47 from packs), 40 (32.26%) milk samples had been contaminated with *E. coli*; 39 (31.45%) from cans and 1 (0.81%) from packs. Resistance was observed in 5 antibiotics out of 11 (45.5%); with the highest resistance to ampicillin (32.5%) and cotrimoxazole (22.5%), followed by ceftriaxone (5%), cefotaxime (5%), and cefuroxime (5%). 2 (5%). *E.coli* isolates expressing extended spectrum beta lactamase (ESBL) were isolated one from pack and one from milk can.

Highest resistance of *E. coli* from the clinical isolates was observed in cotrimoxazole (100%), ceftriaxone (83.3%), cefotaxime (80%), and cefuroxime (75%) followed by ciprofloxacin (69.2%), ampicillin (66.7%), ceftazidime (44.7%), chloramphenicol (27.3%), augmentin (20%), gentamycin (18.2%), and imipenem (14.3%).

The percentage resistance was higher in clinical isolates than in milk isolates for all antibiotics tested. Despite the overall higher levels of resistance observed in clinical *E. coli* isolates compared to raw cow milk samples, there is an interesting relationship between the two sets of results. Specifically, the antibiotics that showed resistance in milk samples also reflected relatively high resistance levels in clinical isolates.

**Conclusion:** The relationship between antibiotic resistance patterns of *E.coli* from raw cow milk and clinical specimens highlights the need for a holistic One Health approach of antibiotic stewardship.

## Introduction

### Background

Worldwide, antimicrobial resistance, especially in food-producing animals has become a serious issue, which requires an immediate intervention [1]. Recent research indicates that food producing animals for example livestock are a reservoir of these resistant bacteria like *E. coli*. These bacteria can transfer in the animal food products such as milk and meat, and are known to cause serious food-borne infections to humans [2]. This has greatly affected the economic status of many countries globally [3].

Morbidity and mortality rates due to bacterial diseases have greatly reduced [4] because of antibiotics globally [5]. However, most of these antibiotics are also administered to animals with either wrong or no prescription at all by continuously adding them to livestock feeds as growth enhancers and promoters [6]. This has led to the rise in the number and strains of bacteria that are resistant to the commonly used antibiotics like extended-spectrum cephalosporins, aminoglycosides such as amikacin, β-lactam antibiotics such as carbapenems and fluoroquinolones like ciprofloxacin [7].

Milk is an animal product that can be fecally-contaminated when there is improper hygiene through animal feeds during milking, storage and handling [8]. Milk is highly nutritious and provides a medium favorable for the growth of bacteria. In fact, it is difficult to attain milk 100% free of bacterial contamination [9]. One method to that can be used to eliminate the contamination is processing the milk. However, only 10 - 20% of the milk in the market is processed, and up to 70% is sold in an unprocessed/raw form [10]. Currently, of the total milk production, only 30% is consumed by farmers and their family members while 70% is marketed. A number of preservation techniques are being used traditionally to store the milk not consumed or for marketing. These include; natural air cooling, traditional vessels like wooden churn and gravitational force [10]. But milk contamination can occur during all these stages of milk production including during transportation to milk shops and also through the cans used in transporting milk to the market and also at the market itself during storage. [3]. Therefore, infection can occur when people ingest the raw milk [3]. Bacteria that contaminates milk includes enteric pathogens such as *E. coli* [10].

*E. coli* is a Gram-negative commensal that resides in the gastrointestinal tract of animals and humans as normal flora [11]. However, some *E. coli* have evolved into different strains of which some are opportunistic pathogens [12], and cause various disease conditions like infections in the urinary tract, intestinal and extra-intestinal infections [6], meningitis, septicemia, peritonitis and also infections in the gastro-intestinal tract as a result of acquiring certain virulence factors [13]. All the pathogenic and commensal strains which have acquired antimicrobial resistance can transfer the resistance to the non-resistant pathogenic and commensal *E. coli* strains in the environment and within animals [14] through horizontal gene transmission [15]. *E. coli* has the ability of easily exchanging its genetic material with other bacteria and harboring a number of resistant determinants. These two factors have made it possible to use *E. coli* as the most common indicator in antimicrobial resistant populations [16]. The emergence, followed by spread of drug-resistant *E. coli* strains is as a result of selective pressure from misuse of antibiotics discovered to treat *E. coli* infections [17].

*E. coli* is one of the most prevalent bacteria found in raw cow milk [18], and this will provide a good underground study of antimicrobial resistance patterns of it in raw milk. There are limited studies that have been carried out in Kawempe Division, Kampala to determine the antimicrobial resistance patterns of *E. coli* in raw milk. This study therefore assessed the resistance profiles in *E. coli* from raw cow milk in comparison with those from clinical specimens obtained from the Clinical Microbiology Laboratory, Makerere University which will be key in identifying the antibiotics to which *E. coli* are most resistant to and hence devising means to reduce and prevent further resistance against effective antibiotics.

## Materials and methods

### Study design and site

A cross-sectional study was conducted from January to March, 2024, in Kawempe Division, Kampala, Uganda on raw cow milk samples from cans and packs collected from milk and on clinical isolates of *E. coli* from the Department of Medical Microbiology, Makerere University, Kampala, Uganda still within Kawempe Division. Kawempe Division is the largest of the 5 divisions in Kampala, Uganda at geographical coordination of 00 23N Longitude and 32 33E Latitude. The population of Kawempe Division is estimated at 388,665 with 19 parishes and 771 villages from which the samples were collected. Written informed consent to provide milk samples was obtained from the milk vendours while permission was obtained to use the clinical isolates of *E. coli*.

### Sample size estimation and sampling

The sample size for the milk samples was determined using the Kish and Leslie method of 1965 where the prevalence used from recent studies [3], a prevalence of 44.57% gave us a sample size of 380 milk samples. However, due to resource constraints, only 124 milk samples were collected. Purposive sampling technique was employed, and only milk samples that did not last for more than two hours at room temperature after collection were included. Random selection of 13 achieved *E. coli* isolates stored between January and March, 2024 from clinical specimens of CSF, urine, blood, sputum, pus aspirates, vaginal swab, and pus swab with age range 14 to 86 years was done. All clinical samples whose demographics were incomplete or mislabeled and damaged milk samples during transportation were excluded. The clinical isolates were used to compare the resistance patterns of *E. coli* with those from milk samples.

### Sample collection and transportation

A total of 124 raw milk samples, each 1 mL were collected from cans and packs from milk shops and in sterile containers from Kawempe Division, Kampala, Uganda. The milk samples were collected aseptically by using methods that prevent contaminations such as gloved hands, sterile wide mouth falcon tubes, and closing the tubes as soon as the milk is collected. The collected milk samples were labeled and packed in a secondary container and transported with an ice box to the Microbiology Department laboratory at Makerere University, Kampala-Uganda with the accompanying laboratory request forms.

### Laboratory procedures

#### Isolation and characterization of *E. coli*

The milk samples were processed immediately for isolation of bacterial growth, identification to species level of *E. coli* and antibiotic susceptibility testing. Briefly, to 1ml of thoroughly mixed raw milk samples 9ml of sterile nutrient broth was aseptically added and incubated overnight at 37°C. The mixture of nutrient broth and raw milk sample was then sub-cultured on sterile MacConkey agar plate under aseptic/sterile conditions and incubated at 37°C overnight for isolation of Gram-negative bacteria and further biochemical tests (TSI, SIM, Citrate and Urease) for confirmation of *E. coli*. The colony units were carefully streaked and inoculated into the TSI, SIM, citrate and urease media and incubated overnight at 37°C. *E. coli* isolates were confirmed according to the CLSI standards and kept in glycerol stocks at -80°C freezer temperature until sub-cultured for antimicrobial characterization.

#### Antimicrobial susceptibility testing

Antimicrobial susceptibility testing was done according to the Kirby-Bauer disk diffusion method to determine the antimicrobial resistance patterns of the *E. coli* isolates following the CLSI protocols (CLSI, 2024.). The principle of the disk-diffusion method for antibiotic susceptibility testing is that antibiotic impregnated discs will be placed on Muller Hinton agar plates which were each streaked with a lawn of the *E.coli* isolate. The plates are incubated for 18-24 hours, and then the diameter of zone of growth inhibition around each disc measured. The readings of the zone diameter were then interpreted as resistant, intermediate or susceptible according to the laboratory standard operating procedures.

Briefly, the *E. coli* isolates were initially sub-cultured from the glycerol stocks and streaked to obtain pure colonies. A sterile wire loop was used to pick colonies. The colonies were emulsified in 4ml of sterile nutrient broth. The turbidity of the suspensions obtained was matched with that of the 0.5 McFarland Standard by comparing the turbidities against a sheet of paper. Using a sterile swab, plates of Mueller Hinton agar was inoculated by lawn streaking; the swabs were evenly be streaked over the surface of the medium in three directions by rotating the plate at approximately 60 to ensure even distribution. With the Petri dish lid in place, the agar was allowed to dry for 5 minutes. Using sterile forceps, appropriate commercially purchased antimicrobial discs were placed evenly on the inoculated plate about 15 mm from the edge of the plate and not closer than about 25 mm from disc to disc. The discs were lightly pressed down to ensure contact with the agar. The antimicrobial discs that were used are in Table 1*. The E. coli* isolates were then classified as susceptible, intermediate, and resistant according to the with the CLSI guidelines of 2024 as illustrated in Table 1

**Table 1.** CLSI standards: Antimicrobial concentrations and interpretation breakpoints for various antibiotic agents used in this study to interpret results.

| ANTIMICROBIAL AGENT<br>(Code) | Disc Drug<br>Concentration<br>(µg) | Breaking Point (mm) |  |  |
| --- | --- | --- | --- | --- |
|  |  | Sensitive<br>(S) | Intermediate<br>(I) | Resistant<br>(R) |
| CRO (Ceftriaxone) | 30 | ≥23 | 20-22 | ≤19 |
| AUG/AMC (Augmentin/<br>Amoxycillin/ Clavulanic Acid) | 20.0/10.0 | ≥18 | 14-17 | ≤13 |
| CTX (Cefotaxime) | 30 | ≥26 | 23-25 | ≤22 |
| CAZ (Ceftazidime) | 30 | ≥21 | 18-20 | ≤17 |
| CXM (Cefuroxime) | 30 | ≥18 | 15-17 | ≤14 |
| AMP (Ampicillin) | 10 | ≥17 | 14-16 | ≤13 |
| CIP (Ciprofloxacin) | 5 | ≥26 | 22-25 | ≤21 |
| IMP (Imipenem) | 10 | ≥23 | 20-22 | ≤19 |
| C (Chloramphenicol) | 30 | ≥18 | 13-17 | ≤12 |
| CN (Gentamicin) | 10 | ≥18 | 15-17 | ≤14 |
| SXT/COT (Cotrimoxazole) | 1.25/23.75 | ≥16 | 11.0-15 | ≤10 |

The table below shows the breaking points (sensitive, intermediate, and resistant) of various antibiotics at particular disc concentrations.

### Controls

Excess fluid obtained during swabbing was removed by pressing and rotating the swab against the side of the tube above the level of the suspension. The *E. coli* ATCC 25922 strains was used as the positive control. After 30 minutes of applying the discs, the plate were inverted and placed in an incubator and incubated aerobically at 35°C for about 18 hours. The control and test plates were examined after an overnight incubation to ensure that the growth is confluent or near confluent. Using a ruler or Vanier caliper, the diameter of each zone of inhibition was measured in mm, on the underside of the plate. Negative controls involved agar plates not inoculated with the any isolates.

### Data management and analysis

The data was entered into; cleaned and coded using Microsoft Excel analyzed using the Statistical Package for Social Sciences (SPSS) version 25.0. MDR and ESBL prevalence was determined using descriptive statistics. A P < 0.05 was regarded as statistically significant.

### Ethical considerations

Ethical approval was obtained from the School of Biomedical Sciences Research and Ethics Committee. Written informed consent to carry out the study from the Clinical Microbiology Laboratory at the College of Health Sciences, Makerere was obtained from the Head of Department of Clinical Microbiology. Written informed consent was obtained from, owners of milk shops and diary milk supply centers and the local leaders in Kawempe Division.

## Results

### *E. coli* prevalence descriptors

This study assessed and evaluated the antimicrobial resistance patterns of *E. coli* isolates from raw cow milk samples obtained from Kawempe Division, Kampala, Uganda, and from clinical specimens at Makerere University, Uganda.

Overall, a total of 124 milk samples were collected (77 from cans and 47 from packs). 40/124 (32.26%) milk samples were found to be contaminated with *E. coli*. Milk samples from cans had a higher prevalence of 39/124 (31.45%) than that from packs 1/124 (0.81%).

The achieved *E. coli* isolates used were obtained from 13 clinical specimens from patients of age range of 14-84 which included blood 2 (15.4%), urine 6 (46.2%). Sputum, vaginal swab, pus swab, and pus aspirate all had 1 (7.7%) sample each.

### Antimicrobial resistance patterns

The patterns of the *E. coli* isolates from the milk samples in this study showed that resistance was higher to commonly used antibiotics. These included Ampicillin AMP (32.5%), Trimethoprim/sulphamethoxazole SXT (22.5%), Ceftriaxone CRO (5%), Cefotaxime CTX (5%), and Cefuroxime CXM (5%) as illustrated in Table 2.

**Table 2:**
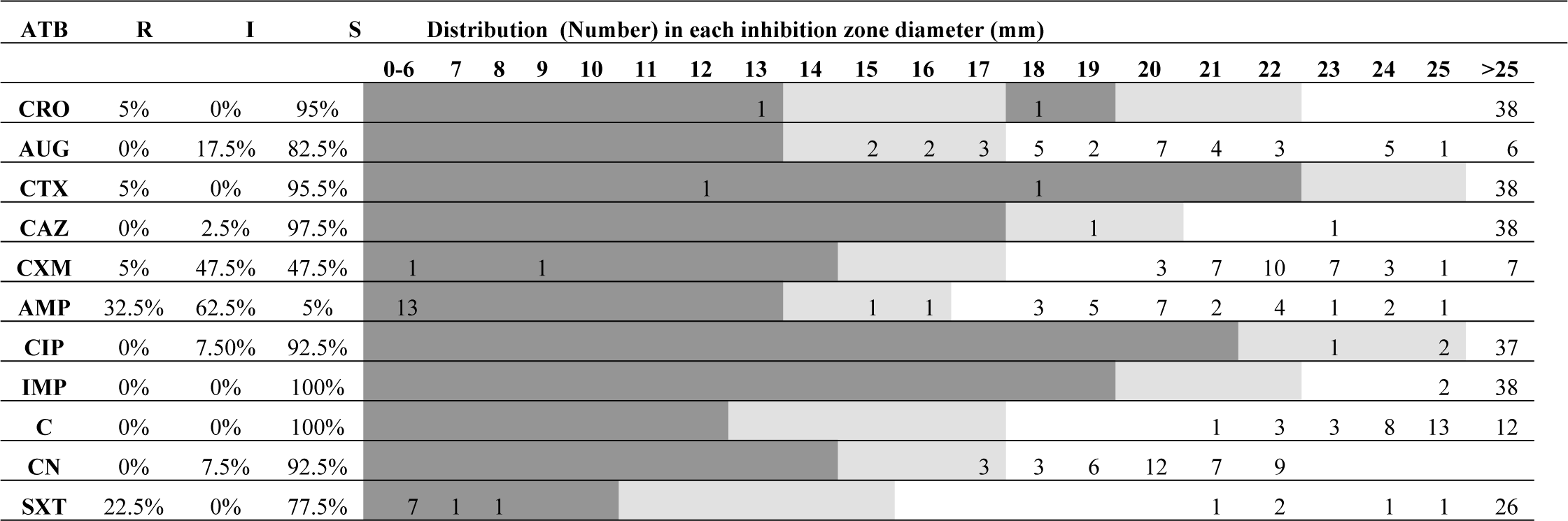
Antimicrobial resistance patterns of *E. coli* from raw cow milk.

The table shows the variation in resistance among 11 selected antibiotics on the isolated *E. coli* isolates from raw cow milk.

The patterns of the *E. coli* isolates from the milk samples in this study showed that resistance was higher to commonly used antibiotics. These included Ampicillin AMP (32.5%), Trimethoprim/sulphamethoxazole SXT (22.5%), Ceftriaxone CRO (5%), Cefotaxime CTX (5%), Cefuroxime CXM (5%).

Highest resistance of *E. coli* from the clinical isolates was observed in cotrimoxazole (100%), ceftriaxone (83.3%), cefotaxime (80%), and cefuroxime (75%) followed by ciprofloxacin (69.2%), ampicillin (66.7%), ceftazidime (41.7%), chloramphenicol (27.3%), augmentin (20%), gentamycin (18.2%), and imipenem (14.3%) as illustrated in table 3.

**Table 3:**
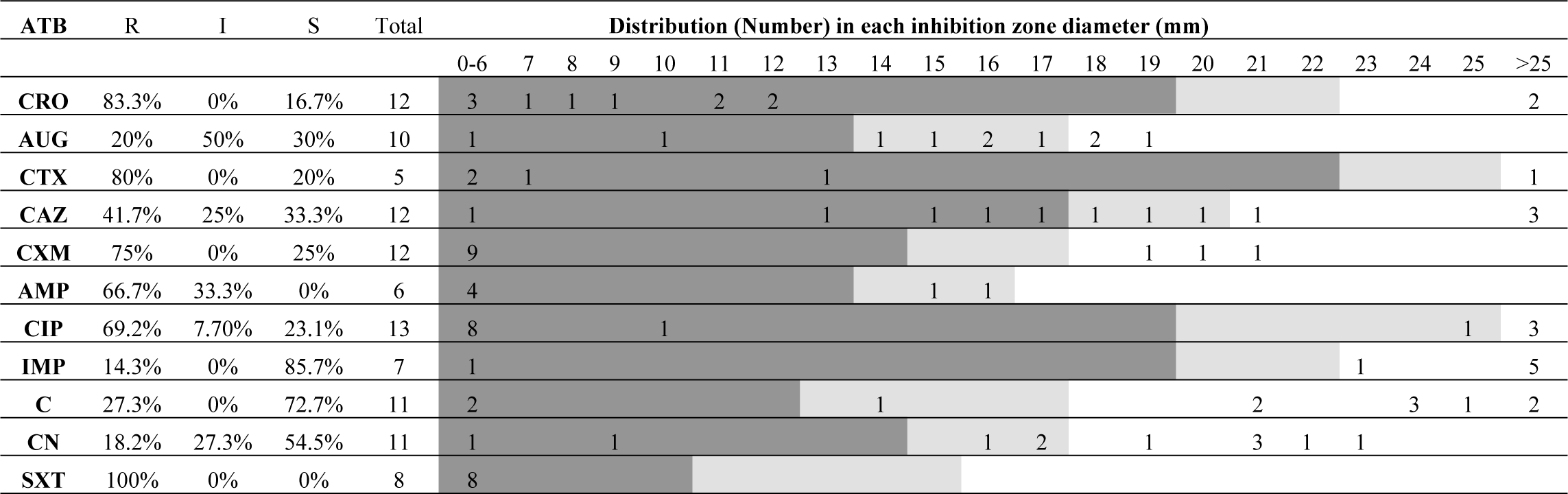
AMR patterns of *E. coli* from clinical isolates.

The table shows the variation in resistance among 11 selected antibiotics on the clinical *E. coli* isolates obtained from the Clinical Microbiology Laboratory of Makerere University, College of Heath Sciences.

The percentage resistance was higher in clinical isolates than in milk isolates for all antibiotics tested. Despite the overall higher levels of resistance observed in clinical *E. coli* isolates compared to raw cow milk samples, there is an interesting relationship between the two sets of results. Specifically, the antibiotics that showed resistance in milk samples also reflected relatively high resistance levels in clinical isolates. This has been illustrated in Figure 1.

**Fig. 1:**
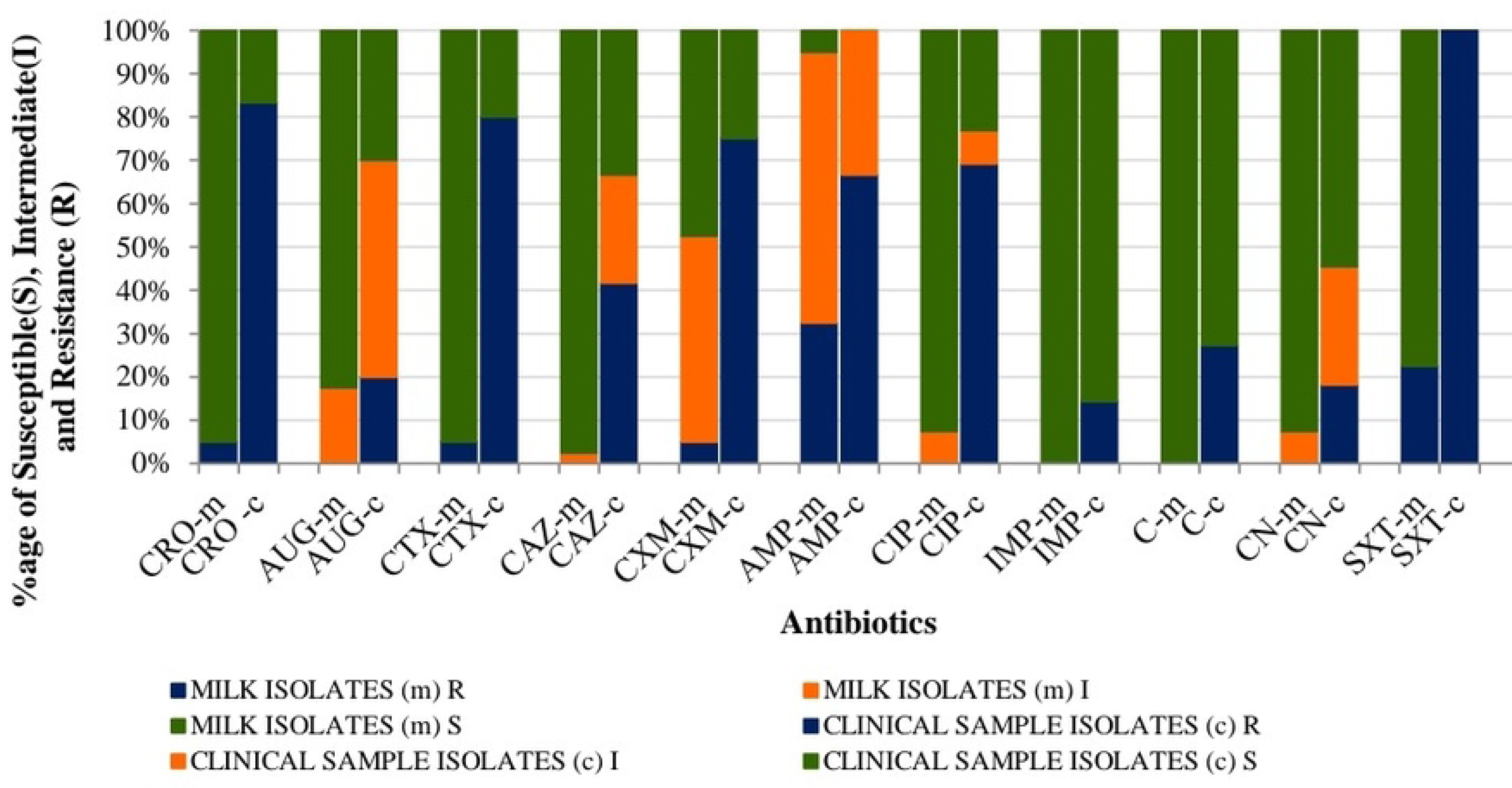
Bar grapgh comparing the resistance patterns of *E. coli* isolated from raw cow milk from Kawempe Divison, and that from clinical specimens from the clinical microbiology laboratory of Makerere University.

The antibiotics used were CRO (Ceftriaxone), AUG/AMC (Augmentin/ Amoxycillin/ Clavulanic Acid), CTX (Cefotaxime), CAZ (Ceftazidime), CXM (Cefuroxime), AMP (Ampicillin), CIP (Ciprofloxacin), IMP (Imipenem), C (Chloramphenicol), CN (Gentamicin) and SXT/COT (Cotrimoxazole).

### ESBL and multidrug resistance

#### *E. coli* isolates from raw cow milk

From the total *E. coli* isolates (n=40 isolates) obtained from the raw cow milk samples, 2/40 (5%) was multidrug resistant at the same time being extended spectrum b-lactamase (ESBL) positive with same AMR pattern of CRO+CTX+CXM+AMP+SXT with resistance to 5 antibiotics as shown in Table 4 and 5.

**Table 4:** Phenotypic patterns of ESBL producing *E. coli* isolated from raw cow milk - 2 isolates.

| Number of antibiotic classes | Resistance patterns | Number of isolates | Prevalence |
| --- | --- | --- | --- |
| 5 | CXT+CRO+CXM+AMP+SXT | 2 | 100% |

**Table 5:**
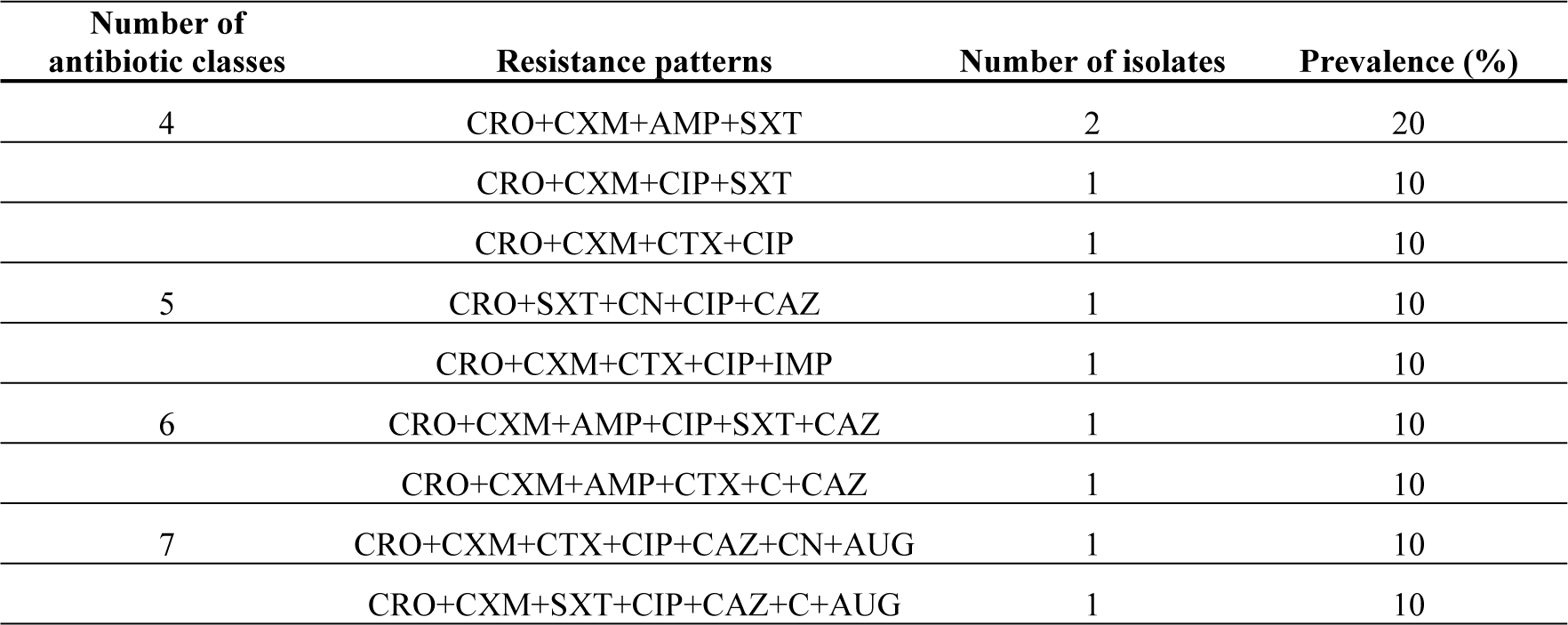
Patterns of antimicrobial resistance phenotypes of *E. coli* strains isolated from Raw Cow Milk.

The table shows the set of antibiotics to which the *E.coli* isolated from milk samples was resistant and attributing to ESBLs.

### The patterns of resistance from this table were determined and used to determine the prevalence for each pattern

#### *E. coli* isolates from clinical samples

Out of the 13 *E. coli* isolates from clinical samples, only 12 were at least resistant to one antibiotic. Of the 12 *E. coli* isolates, the prevalence for ESBL producing *E. coli* was 10/12 (83.3%) while that for the MDR *E. coli* isolates was 9/12 (75%) from the clinical samples. Different phenotypes observed from the 12 *E. coli* isolates were 11 and these showed a great variety of in resistance ranging from 2 to 7 antibiotics. Only the CRO+CXM+AMP+SXT phenotype appeared more than once that is to say two times as illustrated in Table 6 and 7.

**Table 6:**
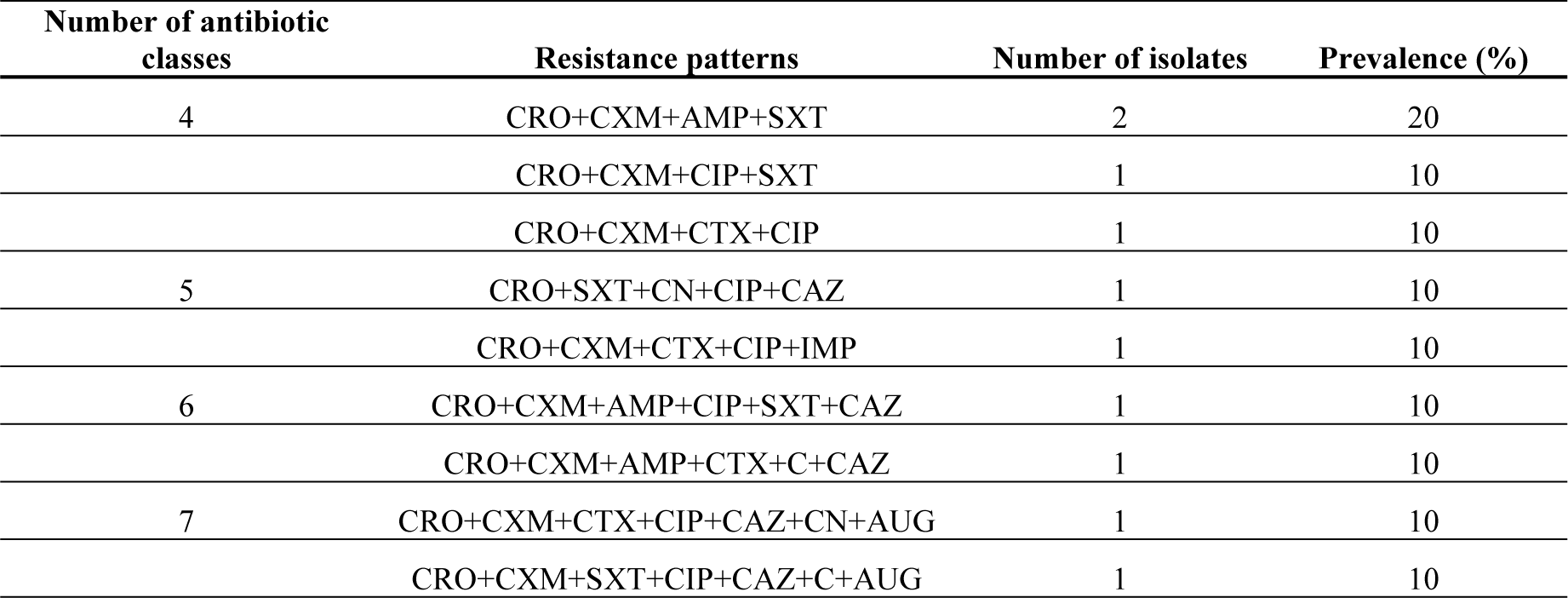
Phenotypic pattern of ESBL producing *E. coli* from clinical isolates - 10 isolates.

**Table 7:**
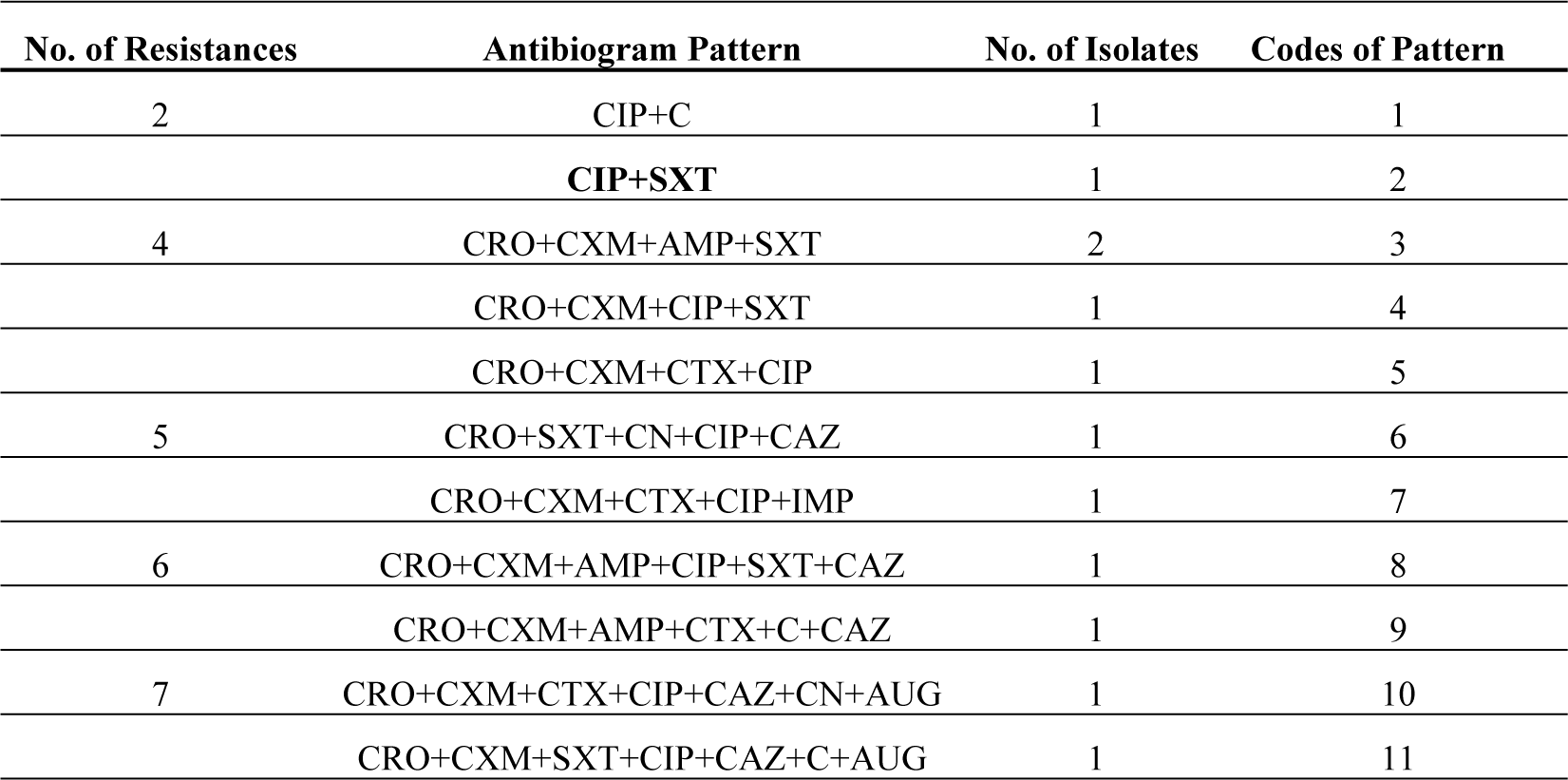
Patterns of antimicrobial resistance phenotypes of *E. coli* strains isolates from clinical specimens in the Clinical Microbiology Laboratory.

The table shows the set of antibiotics to which the *E.coli* isolated from milk samples was resistant and attributing to ESBLs.

The table shows the set of antibiotics to which the *E.coli* isolated from clinical samples was resistant

The proportion ESBL and multidrug resistant *E. coli* isolates was generally greater from clinical specimens than milk specimens.

## Discussion

The rising threat of antimicrobial resistance (AMR) in *Escherichia coli* poses significant challenges to public health, particularly in developing countries. *E. coli* bacteria normally live in the intestines of healthy people and animals. Most types of E. coli are harmless or cause relatively brief diarrhoea. But few strains, such as *E. coli* 0157:H7 can cause severe stomach cramps, bloody diarrhoea and vomiting [20]. This study investigated the prevalence and resistance patterns of *E. coli* isolated from raw cow milk in Kawempe Division, Kampala, Uganda, and compared these findings with those from clinical isolates at Makerere University, Microbiology Clinical Laboratory.

### Prevalence of *E. coli* in raw cow milk

In this study, the prevalence of *Escherichia coli* in 124 raw cow milk samples was 40/124 (32.26%), with a significant difference between cans 39/124 (31.5%) and packs 1/124 (0.81%). This aligns with the study conducted in Ethiopia, which reported a prevalence of 33.8% *E. coli* isolates from raw milk and related products Sarba et al., (2023). Similarly, a study by Torka in 2024 obtained a prevalence of 30.7% of *E. coli* isolated from milk samples collected under poor hygienic conditions Torka, (2024). These results suggest that contamination is widespread in low- and middle-income countries, primarily due to inadequate hygiene practices during handling and storage.The higher contamination rate in milk stored in cans is consistent with a study in Zambia which reported higher prevalence rates of *E. coli* from raw cow milk of 51.2% Mwasinga et al., (2023). This emphasizes the role of environmental exposure and improper sanitation in increasing microbial load. Contrary, the much lower prevalence in packs in this study aligns with findings by Garzon and collegues [24], who highlighted the effectiveness of sealed packaging in reducing contamination.

Globally, *E. coli* prevalence in raw milk varies widely. A higher prevalence of of *E. coli* of 78% in raw bovine milk from North-West of Iran was noted [25], likely due to inconsistent application of hygiene protocols. On the other hand, there was a 30.16% prevalence of *E. coli* in traditional Iranian dairy products. In Nigeria, a prevalence of 44.1% in milk sold in informal markets was observed (Chukwu et al., 2023). These studies corroborate the importance of improved dairy handling practices to mitigate microbial contamination. Regions with stricter hygiene regulations report comparatively lower prevalence rates. For instance, a study done by Dejene et al., observed a prevalence of 6.0 % in central Ethiopia, emphasizing the importance of effective farm-to-market systems (Dejene et al., (2022).

The findings of this study emphasize the need for stringent hygiene measures, especially during milk collection and storage. Regular cleaning and disinfection of milk cans, as well as a shift towards sealed packaging systems, could reduce contamination risks. Such measures, supported by farmer training on proper handling and sanitation, are critical for improving milk safety and public health outcomes (Garzon et al., 2024; Mwasinga et al., 2023).

### Antimicrobial resistance patterns

The antimicrobial resistance (AMR) observed in *Escherichia coli* isolated from raw cow milk and clinical samples in this study reflected alarming trends in AMR prevalence. A significant proportion of raw milk isolates exhibited resistance to common antibiotics, with 32.5% showing resistance to Ampicillin, 22.5% to Trimethoprim/Sulphamethoxazole (SXT), and 5% to Ceftriaxone and Cefotaxime each. In contrast, clinical isolates demonstrated even higher resistance rates: 100% resistance to SXT, 83.3% resistance to Ceftriaxone, and 66.7% resistance to Ampicillin.

These findings are consistent with those of a study conducted in Bangladesh where significant resistance to *E. coli* from milk products was noted with SXT at a prevalence of (33.3%) and Ampicillin at (40%) and also with ampicillin having a higher resistance than SXT [28]. However, the resistance rates were generally higher than those from this study. A study in, Tanzania found high resistance rates in *E. coli* from raw cow milk [29], with highest resistance to Ampicillin at 96.7%, Cefotaxime at 95% Tetracycline 50.4% and SXT at 42.1%. These findings were significantly her than those from this study signifying more antimicrobial misuse hence higher resistance rates. A study by Nayiga et al. Nayiga et al., (2020) also reported overuse of SXT in the treatment of cattle further supporting the high SXT resistance rates. The higher resistance in clinical isolates in our study emphasizes the threat posed by AMR in healthcare settings, as these antibiotics are often first-line treatments for various infections, including urinary tract infections (UTIs) and gastrointestinal infections [31]

The 100% resistance to SXT in clinical isolates is particularly concerning, as SXT is a commonly used antimicrobial agent for treating UTIs [32]. Similar findings have been reported globally; for instance, a study in Pakistan found 78.8% resistance of clinical *E. coli* isolates to SXT [33]. Resistance to third-generation Cephalosporins such as Ceftriaxone in clinical isolates (83.3%) is also worrisome, given its status as a last-resort antibiotic [34]. As reported by [22], the emergence of extended-spectrum beta-lactamase (ESBL) producing *E. coli* strains in both food and clinical samples represents a significant global challenge.

The resistance pattern observed in both raw milk and clinical isolates highlights the potential zoonotic transmission of resistant *E. coli* from agricultural environments to humans. Some studies have shown that *E. coli* resistant to multiple antibiotics in dairy cattle can be transferred to humans through direct contact with animals, consumption of contaminated milk, or environmental contamination (Kasanga et al., 2024a; Tadesse et al., 2018). This route of transmission poses a risk for further dissemination of AMR, especially in regions with high raw milk consumption.

In this study, the prevalence of resistance to commonly used antibiotics underscores the critical need for antimicrobial stewardship in both veterinary and healthcare settings. As emphasized, the overuse and misuse of antibiotics in livestock farming are major contributors to the emergence of AMR, and controlling these practices is essential to combat the spread of resistant pathogens [36]. Integrated AMR surveillance and regulations on antibiotic use are essential to mitigate the risks associated with foodborne transmission of AMR pathogens.

### ESBL and multidrug resistance

Out of the 17 *E. coli* isolates from the 40 milk samples that showed resistance, 2/17 (11.8%) of *E. coli* isolates were ESBL-positive and multidrug-resistant each, compared to 10/12 (83.3%) of clinical isolates that were ESBL-positive, with 9/12 (75%) exhibiting multidrug resistance.

The prevalence of ESBL and MDR positive isolates in milk from this study are way lower than the findings from a study conducted in Indonesia, which reported 7.26% of isolates being multidrug resistant (MDR) and 0.81% of the multidrug resistant isolates being ESBL positive [37].

Another study in Ethiopia documented 65% of the *E. coli* isolates being MDR and 40.8% being ESBL-positive isolates in raw milk [38]. This could possibly suggest overuse and misuse of antibiotics in animals for uses such as growth promoters in these places.

In clinical settings, the higher ESBL prevalence and multidrug resistance reflect significant selective pressure due to antibiotic use. For example, a study in China reported 66.5% being MDR and 73.8% of the MDR was noted to be ESBL positive among clinical isolates, while in Ethiopia, a prevalence of 64.29% multidrug resistance among isolates from clinical settings [39] [39] [39]. This trend corresponds with findings from Georgia et al., 2012 who observed multidrug resistance in 80.4% of clinical isolates, with resistance particularly high to ampicillin (90%), and cotrimoxazole (78%).

In agricultural settings, resistance levels remain relatively lower, although still concerning. [40] found 89.4% multidrug-resistant *E. coli* in raw milk samples from Egypt, with highest resistance to amoxicillin-clavulanate (89.4%), ampicillin (89.4%), cefotaxime (100%) and ceftazidime (100%). Similarly, [22] reported 77% multidrug resistance among *E. coli* isolates from dairy cows in Ethiopia, suggesting that resistant strains in milk could serve as reservoirs for human infections. 7.29% MDR with three different patterns noted [37]

The presence of ESBL-positive strains in milk presents a potential public health risk. A possibility of these organisms entering the food chain, particularly when pasteurization or hygienic practices are inadequate is high [41]. In Ghana, a study reported multidrug-resistant *E. coli* in 57.6% of milk samples and 38.4% of handlers’ hands, underscoring the risk of cross-contamination during handling [42].

The significantly higher ESBL prevalence in clinical isolates reflects the selective pressure exerted by antibiotic use in healthcare. The lower prevalence in milk indicates that while resistant strains are present, and that they may not yet represent a critical threat from the food supply. However, the presence of ESBL-positive strains in milk suggests the potential for these organisms to enter the human food chain, posing a risk for subsequent infections [43]. Continuous surveillance of both food and clinical environments is essential to understand and mitigate the spread of resistant strains through the One Health approach. Recent work emphasizes the importance of monitoring food sources as reservoirs for the resistant strains which can spread through the food chain Despotovic et al., (2023) and [22].

In conclusion, the correlation between resistance patterns in milk and clinical settings emphasizes the need for integrated monitoring and antibiotic stewardship programs in both animals and humans to address the public health implications of AMR.

### Study limitations/recommendations

The sample size of 360 milk samples was not met, as only 124 samples were collected due to limited resources and this could have affected the overall outcome of results. The prevalence of clinical isolates was not determined as already available stored clinical isolates were used. This most likely affected the comparison of the resistance patterns between *E. coli* from clinical samples and those from milk samples.

Also, the number clinical sample *E. coli* isolates (n=13) used for comparison were not equal to of those from milk isolates (n=40) and hence may not have clearly shown the right reflection and relationship as some may not have been from patients from Kawempe division which could have further affected the results. Therefore, a study that is well funded is needed to clearly collect the both samples of milk and human samples from the same study area at the same time.

## Data Availability

N/A. This work is not in any repository as yet

## Conclusion

The relationship between antibiotic resistance patterns of *E. coli* from raw cow milk and clinical specimens highlights the need for a holistic One Health approach of antibiotic stewardship.

## Aknowlegements

I thank the Almighty God who guided me through this season and the wisdom he granted me to develop this idea. I also would like to gratefully acknowledge the support granted to me by my family and friends during the entire process of this research season.

